# Social Cohesion and Covid-19: an integrative review

**DOI:** 10.1101/2023.07.19.23292904

**Authors:** Paul Ware

## Abstract

**Background:** Nations of considerable wealth and sophisticated healthcare infrastructures have seen high rates of illness and death from Covid-19. Others with limited economic means and less developed healthcare infrastructures have achieved much lower burdens. In order to build a full understanding, an appraisal of the contribution of social relationships is necessary. Social cohesion represents a promising conceptual tool.

**Objective:** The aim was to examine scholarship on social cohesion during the Covid-19 pandemic: specifically – the constructions of social cohesion deployed, how it was measured, and the effects of and on social cohesion reported.

**Methods:** The Pubmed, Scopus and JSTOR databases were searched for relevant journal articles and grey literature. 66 studies met the inclusion criteria. Data were extracted and analysed from these using spreadsheet software.

**Results:** Several constructions of social cohesion were found. These concerned interpersonal relationships; sameness and difference; collective action; perceptions/emotions of group members; structures and institutions of governance; local or cultural specificity; and hybrid/multidimensional models. Social cohesion was reported as influential on health outcomes, health behaviours, and resilience and emotional wellbeing; but also that there was some potential for it to drive undesirable outcomes. Scholarship reported increases or decreases in quantitative measures of social cohesion, a temporary ‘rally round the flag’ effect early in the pandemic, the variable impacts of policy on cohesion, and changing interpersonal relationships due to pandemic conditions. There are numerous issues with the literature that reflect the well-documented limitations of popular versions of the social cohesion concept.

**Conclusions:** Social cohesion has been used to express a range of different aspects of relationships during the pandemic. It is said to promote better health outcomes, more engagement with positive health behaviours, and greater resilience and emotional wellbeing. The literature presents a range of ways in which it has been altered by the pandemic conditions.

## Introduction

The Covid-19 pandemic has offered a challenge to those seeking to understand the operation of social relationships and patterns of health and illness. Throughout what is likely the first truly global pandemic in the information age, humans have had access to real-time data from across the globe. What we have seen has frequently surprised us. For example, the 2019 Global Health Security Index positioned the USA and UK in the top two positions in the league table of pandemic preparedness^1^, yet these nations have experienced among the highest rates of illness and death from Covid-19 on the planet^2^. The same report placed Aotearoa-New Zealand (Aotearoa-NZ) and Singapore much lower in the rankings, but these nations have managed to limit their burdens of this kind^2^.

Analyses have sought to explain geographic patterns in populations’ Covid-19 outcomes using, among other independent variables, features of physical geography^3^, previous experiences of infectious disease outbreaks^4^, the ethnic composition of populations^5^ and even leaders’ gender^6^. Although some of these may make important and valid points, none tell the whole story. An understanding of such patterning is not complete without looking to social determinants. After all, it is physical, in-person interaction between humans that allows the virus to spread. In most cases, a nation’s ability to slow or stop the spread of the virus has depended on collective action from the large majority of its population. More fundamentally, the ability of a government to enact policy for protection of public health in the face of Covid-19 is contingent on there being political infrastructure to permit it and, following this, the likelihood that the policy will be adhered to at the population level. At every juncture, the progress of the disease is contingent on social relationships.

One useful tool for the examination of social relationships is the social cohesion concept. Comprehensive accounts of social cohesion have already recently been written^7–9^; however, a brief summary of its history here may provide context to the current discussion. Its inception into academic inquiry is usually attributed to Durkheim^10^, who sought to understand what brings and holds societies together, and occasionally Tönnies^11^. Contributions from Le Bon^12^, Lewin and the field theorists^13^, Parsons^14^ and Lockwood^15^, are frequently acknowledged in the literature. In the 1980s social capital arose adjacent to social cohesion in the works of Bourdieu^16^ and Coleman^17^ and looked more directly at the nature and operation of interpersonal relationships; Putnam^18–20^ later developed his own distinct version. Beginning in the late 1980s, policy scholars and authors took social cohesion for their own and infused it with a range of normative goals for societal development and constructed it in a manner amenable to quantitative measurement. Research institutions in the EU^21–24^, Australia^25, 26^, the UK^27^, Canada^28–30^, and at the World Bank^31^ have all used it as a vehicle toward their own contextually-grounded political aims and analysis. Despite the diversity across constructions of social cohesion through history, disciplinary grounding and politics, it remains, most basically, a tool for the analysis of social relationships and collective sociality.

Given this theoretical lineage of social cohesion and its drawing attention to, among other things, collective action^32^; political or civic engagement^18, 19^; inequality^33^; relationships across population(s), state and social institutions^15, 16^; ethnic and cultural difference^34, 35^; trust^36^; and resilience to hardships or shocks^37^, it appears to offer promise in the quest to understand how social relationships have contributed to different nations’ experiences of Covid-19. This work is an initial step in such a line of inquiry. Beyond this specific topic, and as the direct impacts of the Covid-19 pandemic diminish in magnitude, the accumulation and persistence of further wicked problems – environmental degradation and climate change, poverty and inequality, recurring financial-economic crises etc. – the imperative for understanding collective behaviours becomes ever more salient. Social cohesion is a concept that might be of use to humans’ endeavours therein.

In preparation for an analysis using social cohesion to understand the operation of social relationships within and upon the Covid-19 pandemic, a review of what others have contributed is necessary. Reported here is the process and results of an integrative review on this topic. The integrative method was chosen for its capacity to draw together a range of methodologies^38^. It was of special interest to investigate the different constructions of social cohesion deployed, how they were measured, and the effects of and on social cohesion being claimed.

## Methods

### Search Strategy

A computerised search strategy was conducted on the PubMed, Scopus and JSTOR databases on 10/10/22. Owing to the newness of the topic, keywords used to search for relevant literature were kept simple and broad to maximise results:

> “social cohesion” AND “coronavirus” OR “Covid”

Searches were limited to journal articles and grey literature published since 2020 to exclude those previous to the period of interest. For the PubMed search, the search parameters were set at title and/or abstract. For the Scopus search this was widened to also include keywords, and for the JSTOR search to include the whole text as initial title/abstract searches alone yielded limited results.

From an initial return of 453 results, 56 were reviewed. A further ten articles / reports were included from the reference lists, bringing the total number of papers to 66. Inclusion criteria were:

1. Social cohesion forms a central component of the analysis
2. Discussion on Covid-19 outcomes or social cohesion in the context of the pandemic
3. Full text available through the associated database
4. Full text available in English language

The selection / exclusion process is shown in *Figure 1*.

**Fig. 1:**
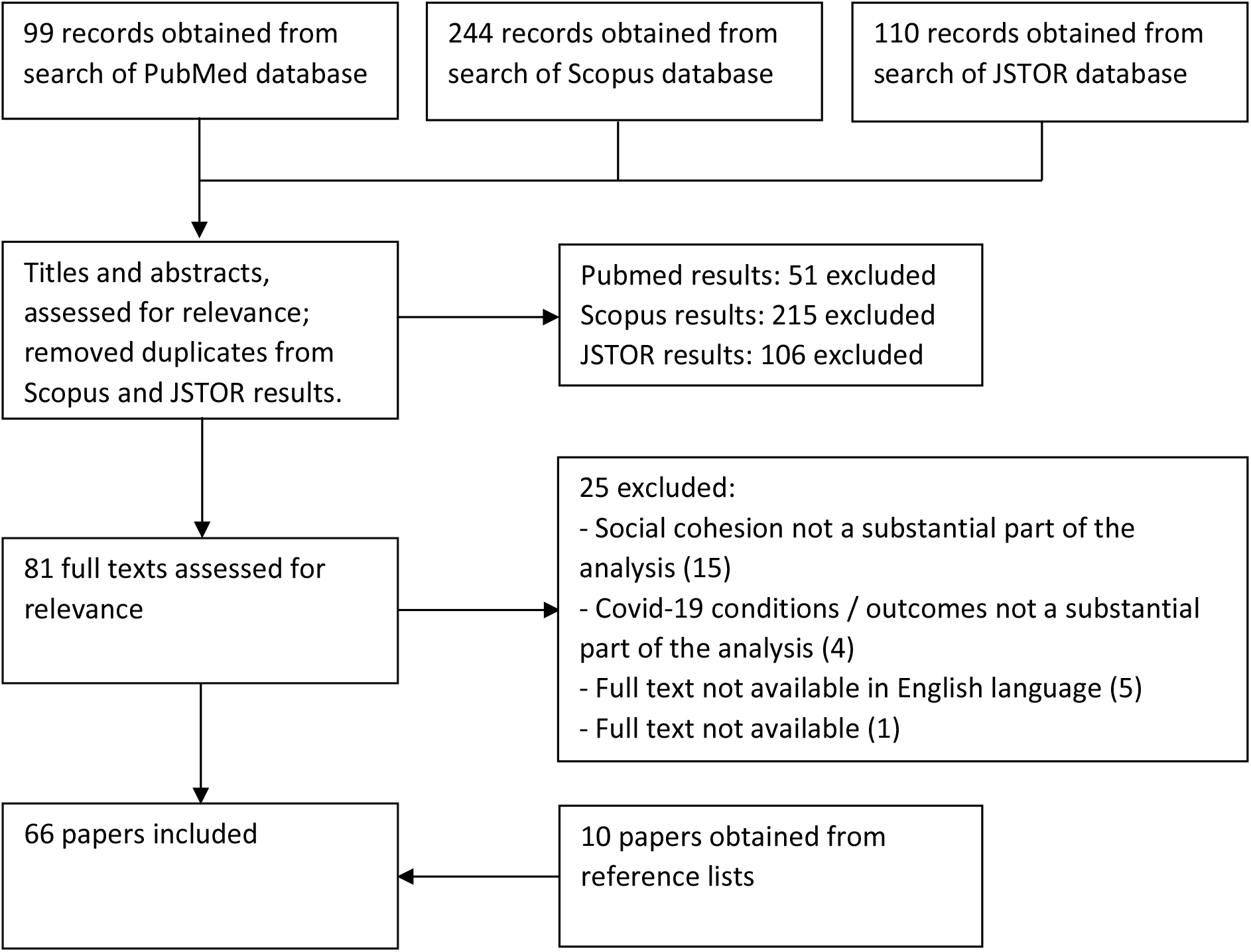
Selection / exclusion process

### Data Extraction

All 66 studies were accessed electronically. In each case PDF files were downloaded from the database on which they were found. Data were extracted using Whittemore and Knaffl’s^38^ method. This consists of data reduction and coding; display; comparison; and the drawing of conclusions. A coding matrix was developed using the Microsoft Excel spreadsheet software (*appendix 1*). During initial coding the literature was arranged by methodology and discipline. Following this, through an inductive process, it was organised by further categories becoming salient during a second pass.

These included the nature of the work (empirical, analytical or theoretical), the unit(s) of analysis, the objects of focus, the construction of social cohesion being deployed, and the outcomes being described. A third pass over the literature informed by this was then conducted, which yielded the thematic structure described in the following section. The extraction focused in particular on the manner in which social cohesion was conceptualised, the components and/or indicators thereof proposed and the argument being presented regarding its operation during the pandemic.

## Results

### Study Features

Sixteen of the pieces of work involved an analysis of two or more nations^39–54^; twelve were based in the United Kingdom^27, 55–65^; seven in the United States of America^66–72^; four each in Germany^73–76^, Australia^77–80^ and Aotearoa-NZ ^81–84^; two each in Iran^85, 86^ and South Africa^87, 88^; and one each in Indonesia^89^, Spain^90^, China^91^, Canada^92^, Argentina^93^, Peru^94^, Denmark^95^, Italy^96^, Kenya^97^, Poland^98^, Romania^99^ and Japan^100^. The remaining three works were not attached to a specific location^101–103^. Fifteen of the papers offered analytical commentaries alone^27, 39–46, 52, 53, 66, 81, 85, 89^ and eight of the papers theoretical commentaries alone^54, 72, 84, 88, 96, 101–103^. Forty of the papers focused their reporting on the collection and/or analysis of empirical data. Twenty-two of these used only quantitative data^47, 48, 51, 56, 58, 65, 67, 69–71, 73, 75, 78, 80, 82, 91, 92, 95, 97–100^, thirteen of these used only qualitative data^49, 50, 55, 63, 64, 68, 77, 79, 83, 86, 87, 90, 94^ and five of these used a combination of the two^59–62, 93^. Three papers combined theoretical and quantitative analysis^57, 74, 76^.

Twenty-eight of the papers’ primary focus was the interplay between pandemic conditions and social cohesion in their location(s) of interest^27, 40, 41, 46, 48, 54, 58–61, 63, 65, 68, 73, 75, 76, 79, 81, 83–85, 87, 88, 90, 95, 98, 101, 102^. Seven of the papers used social cohesion as an independent variable in seeking to explain patterns of health outcomes from Covid-19 between groups^39, 47, 51, 56, 69, 70, 72^. A further seven papers offered discussion on the role of institutions of governance in maintaining social cohesion in relation to the pandemic^42–44, 52, 53, 66, 97^. Eight papers investigated the effects of social cohesion on the practicing of recommended health behaviours or adherence to restrictive public health interventions^49, 50, 55, 57, 71, 80, 86, 100^. Six papers looked into emotional and mental wellbeing in relation to social cohesion during the pandemic^67, 78, 82, 91, 92, 96^. Eight papers presented an analysis of the responsibility of social cohesion for community resilience to the hardships brought by the pandemic^45, 74, 77, 89, 93, 94, 99, 103^. Two papers discussed the effects that the performance of voluntary work during the pandemic had on social cohesion^62, 64^.

### Different Models of Social Cohesion

A range of constructions of social cohesion were presented by the literature. These may be sensibly organised as clusters of concepts and/or objects of attention which are used to represent social cohesion one way or another. These clusters commonly describe the same or similar sets of behaviours or situations using different terminology. Seven are proposed here: one relating to social networks and interpersonal relationships; a second concerning the effects of sameness and difference; a third presenting social cohesion as groups’ collective action or their members’ choosing to act toward the overall benefit of the group; a fourth constructing the concept as a product of the perceptions or emotions of group members; and a fifth concerning the operation of structures and institutions of governance. The sixth category is a small one which contains those suggesting locally and culturally specific expressions of social cohesion, and the seventh consists of accounts presenting large hybrid and multidimensional models containing conceptual representatives of all or most of the other clusters.

#### 1. Interpersonal relationships

Those accounts constructing social cohesion broadly as a product of active interpersonal relationships included those focusing on the provision of social support^41, 55, 65, 68, 71, 77, 83, 90, 94^, interpersonal reciprocity^69, 100^, the sharing of information across social networks^55, 77, 100^, the number of social connections or interactions maintained by an individual^48, 56, 58, 61–63, 65, 72, 74, 89, 93^, the quality of social connections available to an individual^41, 45, 48, 58, 62, 63, 71, 78, 85, 89, 90, 92, 101^ and the relative operation of close relationships such as those between family members and less involved relationships such as those between friends and acquaintances^48, 61–63, 70^. Those operating in this general area frequently referred to a close relationship between social capital and social cohesion^47, 55, 63, 69, 70, 75, 82, 90, 98, 100^, some using these terms interchangeably^93, 94^, or presenting a model of cohesion which more closely resembles what might be considered social capital, usually in Putnam’s^18, 19^ tradition^48, 56–58, 64, 67, 69, 71–73, 77, 78, 85, 89, 91, 92, 101^.

#### 2. Sameness and difference

Where sameness and difference were important to constructions of social cohesion, the axes thereof offered included ethnic groupings^60, 65, 77, 78, 83^, shared identity^27, 60, 61, 80, 97, 102^, norms^91, 102^, values^49, 76, 92^ and behavioural conformity or contagion^50, 55, 76, 77^. For example, Lalot et al.^65^, drawing on Bottoni^104^, present and measure frequency of contact with those of different ethnicity as an indication of greater social cohesion in a society. However, some operationalise ethnic difference in the opposite manner: Morgan et al.^83^ use a model with roots in scholarship from the Canadian Social Cohesion Network^28, 29^ in which the presupposition that ethnic difference presents a challenge for social cohesion is implicit. On a smaller scale, Healey et al.^77^ indicate the importance for social cohesion of having those of similar culture and experience within reach.

#### 3. Working together or acting for the good of the collective

Accounts which emphasised group members’ working together or their choice to act for the good of the collective expressed these behaviours in reference to collective action^39, 42, 43, 52, 55, 65, 66, 79, 80, 87, 99^, co-operation^42, 76, 90, 97, 98^, collaboration^53, 88^, limited conflicts of interest^45^, the pursuit of a shared goal^49^, enactment of social or civic responsibility^27, 47, 50, 55, 87^, a latent orientation to the common good^66, 75, 86^, mutual aid^45, 85^, altruism^66, 78^ and solidarity^45, 46, 49, 50, 66, 91, 103^. In this respect, Garber and Vinetz^66^ consider the tension between adjacent processes of individualised identity and low trust in public officials, and the importance of cohesion as collective buy-in for public health interventions such as mask wearing.

#### 4. Social cohesion as a subjective phenomenon

Some accounts presented social cohesion as the sum of individuals’ appraisals of the group or population of which they were a member. Orientations measured included feelings of togetherness or unity^27, 50, 60, 76, 79^; trust in hypothetical others from the same nation^47, 61, 63, 73–75, 80, 90^, or those in one’s neighbourhood or community^39, 47, 56, 58, 60, 61, 67, 69, 74, 75, 78, 80, 85, 86, 90–92, 101^; trust in others to follow public health advice and/or adhere to formal restrictions^59–61, 76^; trust or confidence in public officials, politicians or political institutions^39, 47, 60–63, 65, 66, 74, 76, 80, 87, 90, 93, 95, 97^; or, simply, perceived cohesion^58, 86^. For example, Collischon and Patzina^73^ place trust of ‘unknown others’ at the centre of social cohesion and find geographically patterned negative association with Covid-19 incidence, an effect mostly driven by women in their sample, hypothesising that this may be driven by the suspicion that others’ neglect to follow public health advice.

#### 5. The operation of institutions and structures of governance

Some accounts of social cohesion situated it at the structural level. Such accounts included those that related social cohesion to the effectiveness of democracy in a nation ^51^; the integrity of the social contract^44^; within-nation inequality^43, 44, 47^; governments’ action to uphold human rights^95, 103^; the strength of state institutions^49, 53^; coherence and integration of institutions, state tiers or other structural elements (Lockwood’s^15^ ‘system integration’)^54, 86, 97^; a state’s tactics and success in relation to internal conflict management and maintenance of order^103^; and effective leadership^43, 53, 102^. Razavi et al.^44^ conducted an analysis using such a version of social cohesion, suggesting that pre-pandemic declines thereof in European nations was temporarily arrested by governments’ instituting welfare policies to ameliorate the economic disruption of stay-at-home orders.

#### 6. Locally or culturally specific arrangements of cohesion

A small number of commentators suggested that there are distinct and locally specific arrangements of social cohesion. Schröder et al.^76^ described a latent network of connections across local populations as a ‘milieu’ from which specific forms of cohesion arise in relation to the arrangements of culture and socioeconomics within. Villalonga-Olives et al.^90^ gave an account of the culturally bound manners of relating in the Mediterranean region and Spain. Garber and Vinetz^66^ noted the culturally specific forms of resistance and their place in questions of social cohesion more broadly that lead to patterns of rejection of restrictive public health measures^66^. Miao^91^ made comment on the specific cultural-historical tendency of Chinese peoples to act for communal benefit, this implying a distinct operation of social cohesion in their population of interest. Saghin^99^ found that the localised tension between a burgeoning culture of individualism and the hangover from communist collectivism was instrumental to social cohesion contributing to Romanians’ resilience during the early pandemic. Metz^88^ discussed the African ethic of *Ubuntu* (an expression of collective orientation), how this creates a locally specific form of social cohesion, and how those existing within this social environment might negotiate the demands of distancing during the pandemic.

#### 7. Hybrid and multidimensional models

There was widespread (but not ubiquitous) recognition in the literature that social cohesion is usually understood as a complex and multidimensional model. After acknowledging this fact, most works included in this review proceeded to focus on, analyse and/or measure one specific concept or cluster. However, some authors sought to deploy a broad focus facilitated by a systematically integrated collection of concepts. Among these, some referenced pre-existing theoretical models assembled by other authors^65, 83^, some detailed the construction of their own theoretical models for the specific purpose of the work being reported^27, 76, 103^, and others used different combinations of conceptual categories to represent social cohesion, using a range of justifications for their choices, but stopping short of building these into a free-standing structure^49, 80, 97, 103^. Of the two works in the first group, one^65^ used Bottoni’s^104^ model, which incorporates appraisal of interpersonal relationships, collective action and the operation of the state. The other^83^ used a model initially constructed by the Canadian Social Cohesion Network^29^ then later developed and given Aotearoa-Aotearoa-NZ specificity by Spoonley et al.^105^. This model appraises sameness / difference, collective action and the operation of the state. Of those in the second group, one^27^ offered a model constructed from concepts relating to sameness and difference, collective action or working for the good of the collective, and subjective processes. Another^76^ following the construction of a novel theoretical model, sought to represent social cohesion in the context of the pandemic in Germany in relation to subjective appraisals of togetherness and the operation of structures of governance. The third^103^ situated social cohesion as a component of a much larger model of resilience during crises. In this model, social cohesion was constructed using concepts relating to interpersonal relationships, subjective appraisals, collective action and the operation of the state. The remainder offered combinations of subjective appraisals, collective action and the operation of the state^97^; subjective appraisals, interpersonal relationships and the operation of the state^74^; collective action, sameness and difference and the operation of the state ^49^; and sameness, subjective appraisals and collective action^80^.

### Units of analysis

There were a range of different units of analysis across the papers reviewed, largely contingent on the manner in which social cohesion was constructed or measured by each. Most commonly, characteristics of collections of individuals were observed in order to describe populations at the local geographic level^45, 56–58, 61, 64–67, 69, 71, 72, 77, 78, 82, 85, 86, 89, 91–93, 96, 99, 100^. However communities of sameness ^94, 101^; regions within nations ^70^; entire national populations^40, 88, 95, 97, 98^; and nation states themselves^39, 42–44, 46, 47, 49–53^ were also used as the units of analysis. National populations were often divided along demographic axes in order to analyse specific groups: occupational^62^, age^41, 83^, gender^73^, or a combination of these axes of difference^81^. Some – often the qualitative work and that coming from researchers in psychological fields – focused on individuals^48, 55, 63, 68, 74, 79, 103^. Finally, a number of works used some combination of the above units of analyses: individuals and social groups^87, 90, 102^; neighbourhood and social groups^76^; communities of sameness and social groups^54^; neighbourhoods and nations^80^; neighbourhoods, regions and nations^27, 75^; individuals, neighbourhoods, regions and nations^59^; and individuals, social groups, communities of sameness and regions^60^.

### The different effects of social cohesion

#### 1. Health outcomes

Those works presenting social cohesion as a determinant of health most commonly held it to be producing comparatively positive population health outcomes from Covid-19. Such reporting included the negative association of social cohesion or social capital with the likelihood of death^51, 70^; or the spread of infections of Covid-19 in groups and populations^69, 72^; or antibody response to vaccination^56^; or, in the case of some analytical and theoretical work, a less precisely defined and more general ‘health outcomes’^39^. For example, Gallagher et al.^56^ used a 5-item questionnaire to measure frequency of contact with neighbours; and perceptions of neighbourhood trustworthiness, willingness to help, similarity and friendship; and vaccination-related blood antibody concentration among 676 people from the UK, finding a positive association between their measure of social cohesion and blood antibody concentration.

Some works sought to isolate the effect of different components or variables within their model of social cohesion on health outcomes from Covid-19^47, 69, 70^ and/or different effects of the same components on different populations^69^. For example, Elgar et al.^47^ drew on existing data from surveys across 84 countries which used a Putnam-inspired construct measuring trust in other people, membership of community groups, civic activity and confidence in the state, to investigate a hypothesised association between social cohesion / capital and Covid-19 deaths in the early days of the pandemic. They reported that while mortality was positively associated with interpersonal trust and group affiliations, it was negatively associated with civic engagement and confidence in the state. Ransome et al.^69^ measured feelings of belonginess, trust in neighbours, perceptions of neighbours’ willingness to help, civic and social participation and collective engagement across a selection of neighbourhoods in Philadelphia, looking for any associations with rates of Covid-19 diagnosis. They found that social cohesion operated differently in different places: neighbourhoods predominantly occupied by African Americans found some of their indicators of choice to be associated with higher rates of diagnosis, whilst those in which African Americans were the minority saw the same indicators associated with lower rates.

#### 2. Information for, and practice of, health behaviours for the prevention of Covid-19

A number of works described the effects of their chosen construction of social cohesion on the promotion of health behaviours intended to prevent transmission of SARS-CoV-2. These included commentary on those behaviours made compulsory by legislation and access to or transmission of information around them. This body of work includes notable contributions from those who deployed qualitative methods to explore in detail the motivators and barriers for practicing recommended and required public health behaviours^49, 50, 55, 86^. Each of these introduced social cohesion at the latter stages of their analyses to draw together a collection of influences on opinions and behaviours into a single explanation. In keeping with the qualitative approach, these presented a more complex arrangement of forces than the one-dimensional scales or binaries commonly expressed by quantitative work. For example, Zimmerman et al.^50^ used social cohesion to explain those behaviours informed by feelings of togetherness, commonality, empathy and compassion; linking this to the spread of uptake of health promoting behaviours across a group. They noted that these can encourage closer adherence to public health guidelines through care for the collective, while at the same time may cause people to deviate from guidelines and regulations when doing so is perceived to benefit another or others who may be in need.

Others produced quantitative analyses using self-report surveys to capture the practice of specific sets of Covid-19 related health behaviours^71, 80, 93^. For example, Cheng and Lo^71^ surveyed older adults from the USA on the number of preventative behaviours from a given list in which they engaged and a 3-item Likert scale measuring neighbourhood social cohesion. They reported a positive association between the two. Cardenás et al.^80^ surveyed a large sample of Australians to investigate whether engaging in physical distancing and hand hygiene behaviours were susceptible to socio-political determinants. They used a model of social cohesion incorporating social identification, confidence in government and social relations captured by a 14-item tool. They reported a complex set of findings in which their measures relating to social cohesion are spread across being positively associated with these health behaviours, negatively associated with them, evidencing no apparent relationship, or associated with one and not the other.

The place of social cohesion in the distribution, provision and accessing of good quality health-related information has also been of interest to scholars^55, 77, 100^. Burton et al.’s^55^ and Healey’s^77^ interview research presented the sharing of health information as a form of social support and this, in turn, as a component of social cohesion. Machida et al.^100^ suggested that the offering and uptake of health information between groups bound by ethnic and cultural sameness may be an explanation for the positive relationship they found between social cohesion and rates of vaccination.

Some authors engaged in commentary on relationships between social cohesion and whether or not people choose to become vaccinated against Covid-19^57, 100^. For example, Machida et al.^100^ in a cross-sectional study in Japan, found social capital (in their Putnam-derived construction containing social cohesion) to be associated with both previous vaccination and intent to receive a booster vaccine.

#### 3. Social cohesion promoting resilience and emotional wellbeing

The effects of social cohesion on resilience and/or mental and emotional wellbeing have seen significant attention in the literature. These works commonly noted the heightened states of stress and anxiety brought about by the pandemic and the isolating effects of the physical distancing measures that were employed to prevent transmission of Covid-19. There are those who referred to the resilience of groups in this respect, proposing that their chosen model of social cohesion is supportive of this^45, 77, 89, 93, 94, 99^. For example, Rela et al.^89^ offered an analysis of the action of a Putnam-influenced fusion of social cohesion and social capital in Indonesia. An argument is presented for social cohesion as a determinant of a community resilience, defined as the capacity of a group to cope with changing environments while continuing to improve the living conditions of its members. Garcia-Rabines et al.^94^ and Healey et al.^77^ both argued that within-group cohesion in marginalised communities (trans women and ethnic minorities respectively) can act as a source of resilience in times of crisis in the forms of practical and moral social support. Group resilience is also, by some, explicitly held as important for groups’ protection against psychosocial distress and clinical versions of this^74, 93^. Most commonly, however, the various constructions of social cohesion at the group level are offered as a resource for individuals’ protection from poor mental health outcomes^67, 78, 82, 91, 92, 96, 103^. For example, Best et al. ^92^ surveyed 1,381 Canadians on perceptions of social cohesion in their neighbourhoods and levels of panic, depression, emotional stability and worry, alongside a range of other variables. They found that the pandemic and the restrictive responses to it were responsible for heightened distress, but that their social cohesion indicators were unequivocally negatively associated with all measures thereof. O’Donnell^78^ reported, on the basis of longitudinal research with Australian informants, that neighbourhood-level social cohesion was associated with lower levels of depression during periods of high infection rates and restrictions on social activity; but that there was no effect on anxiety and loneliness.

#### 4. Disunity, fragmentation and the negative effects of social cohesion

Some authors offered commentary on the opposites of social cohesion, or its less desirable effects. A number of works presented social cohesion and division as antithetical and presented broad commentaries on the operation of the latter^27, 43, 54, 60, 61^. For example, Bisiada^54^, in a theoretical discussion of social life in Germany and Spain over the course of the pandemic countered the argument that social divisions have been primarily ideological in nature and presented another, that socioeconomics have been the major axis. Abrams et al.^60, 61^ presented data to show that the United Kingdom’s experiences of the pandemic have been marked by increasing perceptions of unity at the local level but of disunity at the national level. Other works considered the manners in which social cohesion as it is constructed may produce undesirable outcomes. These include social cohesion or elements of it as a driver of increased infection or death rates. Ransome et al.^69^, using data collected from across the USA, and Thomas et al.^72^, in a San Francisco based simulation, suggested that social cohesion viewed as collective engagement or frequency of interpersonal connection respectively may offer a mechanism by which increases in social cohesion may explain ethnic differences in infection rates. Brief commentary was also made by Hangel et al.^49^, Zimmermann et al.^50^ and Schneiders et al.^63^ on the manner in which demands made by commitments to the cohesion of a larger social group might come to negatively impact the wellbeing of those in one’s close circle. One example of this is in the demands for physical distancing in order to stop the population-level spread of SARS-CoV-2 meaning that some family members suffer isolation.

Some works presented commentaries suggesting that social cohesion may operate to produce in-group versus out-group orientations or sharpen existing ones in populations in the pandemic environment and explored some of the implications^42, 81, 83, 90, 95^. Comment was made on group constructions that pre-existed the pandemic and their boundaries and operation in its context: age^83^, ethnicity^83^, gender^42^, and neighbours versus ‘outsiders’^90^. Others suggested that new groups have formed in response to the pandemic and the requirements of the public health response^81, 95^.

Ergler^81^ offered an analysis of the procession of groups that have become marked as outsiders as a result of the Aotearoa-NZ pandemic response which included political efforts to generate a national unity in opposition to the virus. Schuessler^95^ measured the extent to which compulsory vaccination policy in Denmark brought into being group boundaries which exclude those resisting vaccination from some aspects of social life.

### Changing social cohesion during the pandemic

#### 1. ***‘***Increasing’ and ‘decreasing’ cohesion during the pandemic

Some commentators suggested that the pandemic conditions facilitated increases in social cohesion in the manner they constructed it, that is, the quantitative measure(s) on which the appraisal is built. These suggestions were based on observations of increases in the provision of social support^68, 79, 83^; the fruitful interaction between pandemic conditions and previous policy efforts to bolster social cohesion at the local level^65^; and perceptions of increases in local unity and solidarity^27, 60, 61^. For example, Morgan et al.^83^, conducted in-depth interviews with older people following the first wave of the pandemic in Aotearoa-NZ and found participants feeling a greater sense of belonging in their communities due to the help they received from family and friends during a difficult time, this being experienced in different manners by different ethnic and age groups. Some (including some of the same authors) also painted the opposite picture: the pandemic environment bringing about a reduction in social cohesion. Such evidence presented includes declining trust: in others from the population under investigation ^65^, and in politicians and political institutions^60, 61^. Perceptions of declining national unity are also referenced in this respect^27, 60, 61^. Others, as covered above, reference the action of policy responses to the pandemic to create division^74, 81, 95^; and the occasional failure of national policy approaches to cater to and include all groups^83^. Also raised in this respect is the sometimes polarising effects of pandemic politics and public health responses and the interplay thereof with social cohesion, adherence to public health measures and population health outcomes^51, 54, 95, 98^.

#### 2. *‘*Rally round the flag’ and its diminishing returns

An observation that is made frequently across the literature, especially in works that have engaged in longitudinal research or ongoing analysis, is that there was a moment of increased cohesion in the early stages of the pandemic which gave way to a return to form – or worse – toward the end of 2020. This was characterised by the British Academy^27^ as a “rally round the flag” effect. This effect has been described occurring in relation to a range of different constructions of social cohesion and their indicators: interpersonal interaction, social support and feelings of togetherness^27, 68, 87^; trust in ‘most people’^73^, those in one’s neighbourhood^59^ and government^27, 60, 93^; political polarisation^98^; feelings of national and local unity^27, 59–61^; and collaboration between nations^46^. The British Academy’s^27^ UK-based narrative suggested pre-existing division and declining cohesion in the years before the pandemic, followed by a ‘coming together’ during the first wave, in which people were brought to trust each other more, have increased perceptions of local and national unity, identify more closely with their communities and offer both emotional and practical support to others. The authors suggested that this moment had come to an end by September 2020 at which point almost all indicators returned to pre-pandemic levels. Perceptions of local unity in some cases remained higher, and trust in government declined to still lower levels, the latter effect being most pronounced in socioeconomically deprived communities and ‘key’ workers such as those employed in front-line social care, who were concentrated in these lower socioeconomic strata, and who experienced the greatest exposure to risk of contracting the virus.

#### 3. The effects of policy

A smaller body of work looked into the effects of policy on social cohesion in pandemic conditions. There have been those already mentioned who held vaccination policies^95^ and a framing of collective action against the virus^81^ as responsible for driving division. Others pointed to the greater levels of social cohesion experienced by nations who focused policy towards fostering it in their pandemic response – including those emphasising solidarity and agency^42^; “we-ness”, that is, feelings of unity^43^; and the swift roll-out of generous social protections ^44^. Contrary to this, Strupat^97^, in an analysis of the Kenyan response, considered that the burden placed on the nation by the pandemic was too great for the welfare approach overseen by the government to have any real effect. A further set of commentaries, all from the early stages of the pandemic, offered analyses and advice for policy authors toward prioritising social cohesion in the pandemic response^45, 52, 53, 66^.

#### 4. Changing interpersonal relationships

A significant thread in the literature, involving those constructions of social cohesion where interpersonal relationships are at the centre, is analysis of how changes in relationships brought about by the pandemic have affected social cohesion. Such accounts include the suggestions that frequency of contact and quality of relationships with people from outside the family circle have diminished^48, 58, 60, 61, 75, 85^; and that bonding with those in one’s close environment has strengthened^48, 59–61, 63, 83^. Other interesting observations on the changing nature of interpersonal relationships includes their becoming politically polarised and/or depolarised over time^63, 98^; and the manners in which the requirements of remaining physically distant can function in cultures of communal sociality^40, 88^.

A common suggestion is that the conditions of the pandemic have produced a fundamental reformation of the manners and practices by which social relationships are enacted – and therefore a change in a cohesion which is reliant on these. These range from, most simply, the observation that the novel social conditions of the pandemic may bring new conditions for the construction of personal identity, group identity and thus group interactions^102^, that the extent of disruption to established practices has required a conscious and intentional reformation of relationships^63^, to the idea that times of crisis bring the requirement for specific sets of practices of interaction^76, 78, 94^.

More specific observations include those on details of the large-scale shift of interaction to remote communication technologies^68, 79^; the use of outdoor singing as a communal negotiation of grief and solidarity^96, 101^; and signing up to volunteer programmes as an expression of commitment to the collective and a means of engaging in meaningful interpersonal interaction^62, 64, 75^. There are numerous commentaries dealing with the changing practices of interaction of different groups, including the changing construction of oldness in relation to older people’s increased vulnerability during the pandemic and how this has affected inter-generational relations^41, 79, 83^; the ‘closing ranks’ of trans people in expectation of heightened oppression^94^; increasing victimisation experienced by ethnic minorities^60^; the increase in disconnection and isolation felt by key workers ^62^; and the disproportionate decline in quality and frequency of social relations felt by socioeconomically marginalised peoples^27, 43, 44, 54^ and those of lower formal education levels^75^.

Of particular interest due to their novel nature are the accounts that identify the intersection between local culture and pandemic environment to create conditions of possibility for new practices of interaction. Schröder et al.^76^, upon their aforementioned theoretical model in which cohesion arises in a space of latency from an underlying milieu of social relations, noted that in Germany these practices took different forms in different geo-cultural regions. These authors held the distribution of socioeconomic resources and values to be of special importance. Villalonga-Olives et al.^90^ described a ‘bounded solidarity’ on the island of Menorca in which there was an increase in solidarity and support in the communities of the island but a tension between mistrust of outsiders as potential carriers of disease and the reliance of the same outsiders to bring in capital to maintain islanders’ financial wellbeing.

### Critique

The literature on social cohesion and the Covid-19 pandemic contains weaknesses. A large proportion of these limitations are a function of the already well-documented issues with the social cohesion concept. The broad diversity of definitions and constructions of social cohesion have long been the subject of discussion and appear to be no closer to a resolution^7, 8, 106^. In respect of the literature under analysis here, this means that there is such a spread of versions of social cohesion in circulation that its explanatory power is weakened and it is difficult to make stable comparisons across the body of knowledge. In general, Bernard’s^33^ well-quoted observation remains apt: social cohesion is a “quasi-concept”, amenable to the imprinting of any political-ideological or discipline-bound framework within which those using it are working. The presence of such diversity within social cohesion and its value-laden nature necessitates explicit theoretical engagement and clear justification of choice for the version of the concept being deployed. With some notable exceptions (e.g.^76, 83^), this work is not – beyond an acknowledgement of diversity – undertaken with detailed attention in the literature reviewed here. As such, social cohesion does not carry as much explanatory strength in analyses of the conditions of the pandemic as it might: a model built without full attention to theoretical foundations leaves its conclusions open to concomitant critique.

In this body of literature, social cohesion is, as mentioned, situated among a group of concepts clustered around a set of behaviours, orientations and situations. Social cohesion is frequently not precisely defined or distinguished from those concepts adjacent to it. This is especially true in respect of social capital where the two are often treated as synonymous (e.g.^77, 89^) or where social capital is used as a proxy for social cohesion without a detailed justification (e.g.^57, 93^). There is also frequent unacknowledged definitional imprecision across social cohesion and ‘solidarity’ (e.g.^49, 66^), engagement with democratic processes (e.g.^51^), the social contract (e.g.^44, 73^) and activity in the civic space (e.g.^99, 100^). Where these concepts are not defined and differentiated with care and clarity, analyses become unclear as to the networks of cause and effect being invoked and, again, explanatory power is impeded.

It is possible that a large portion of the imprecision across social cohesion, social capital and civic engagement is a consequence of the reliance on Putnam’s^18, 19^ framework. This model, nominally of social capital, incorporates several diverse elements. It situates social cohesion in part as a product of interpersonal connections and individuals’ access to resources (finance, support etc.); a notion present in other constructions of social capital^16, 17^. It also, however, measures political and civic engagement and uses whole groups as the units of analysis – practices which might sit more comfortably under the social cohesion banner. This may invite conflation of these concepts. The frequent use of Putnam’s framework also means that the body of knowledge focusing on social cohesion during the pandemic is heavily skewed toward a cluster of constructions of social cohesion kept inside its boundaries: at the local/neighbourhood level, contingent on its specific set of indicators and its knowledge produced by quantitative inquiry. One common consequence of this is a self-fulfilling deficit-orientation by which social cohesion is held to be reliant on face-to-face interpersonal interaction and trust, is measured quantitatively with models designed for normal conditions in the context of stay-at-home and physical distancing requirements, and it is inevitably concluded that there are problems for social cohesion as a result.

An over-reliance on quantitative models is accompanied by an over-reliance on old established measures of social cohesion from self-report survey data to impede the quality of knowledge in the area. Although the complexity and local specificity of social relationships and the unprecedented nature of the pandemic environment are well recognised, the measurement of social cohesion using, for example, Likert scales with five or fewer items – sometimes one alone – is common and is unlikely to be sufficient to describe the complexity of the relationships across groups and the populations under scrutiny in novel circumstances. One such frequently deployed tool is Sampson’s^107^ scale. This was devised in 1997 to measure disorder and collective efficacy in the suburbs of Chicago and measures a number of perceptions of the neighbourhood on a 5-item Likert scale. Also common practice is to ask a single question on perceptions of other’s trustworthiness.

Aside from the potential problems with validity in the novel pandemic context, the problems with making firm conclusions on self-report data of this kind are well-documented (e.g.^108^). Even when cultural specificity and the newness of the pandemic environment is explicitly acknowledged, the complexity and diversity of social relations are recognised, and the ongoing issues with the social cohesion concept engaged with, some still fall back on such limited manners of measurement^76^.

At the other end of the spectrum, there are some models which appear to be so large and all-encompassing that social cohesion becomes the basic determinant of almost all psycho-social life and culture. For example, Godara et al.’s^103^ construction includes social connections (capital), interaction, inclusion, civic engagement, identity, social structures, norms and values, loyalty, solidarity, human rights, trust, conflict management, equality and order; across micro, meso and macro levels; involves both structures and groups of various sizes; and includes vertical and horizontal relationships. Scholarship of this kind creates a version of social cohesion that becomes overly mechanistic, un-attentive to local difference; while at the same time too large to make work within the limits of the capacities of knowledge-production apparatus. Its broadness also, again, weakens its explanatory power, such that Schröder et al.^76^ warn that the concept is in danger of becoming an “empty signifier”. There may be a sweet spot between reductionism, expansionism and the particular that few in this body of knowledge have managed to achieve, though there are some notable examples, predominantly using qualitative or mixed-methods (e.g.^27, 60, 83, 90^).

Two further problems with the literature reviewed here are also noted by commentators on the social cohesion literature more generally. The first, in relation to the quantitative work, and described previously by Janmaat^109^ and Green and Janmaat^110^, is that the indicators contained within commonly deployed constructions of cohesion frequently do not co-vary (e.g.^80^). This calls into question the validity of the construction of a concept which aims to provide a single coherent explanation – or independent variable – for social life. The second is that there is some ethical concern with works^69, 72^ that apply closed-ended quantitative approaches to identify ways in which social cohesion may be driving higher rates of sickness and death from Covid-19 in marginalised populations and minorities. As has been pointed out elsewhere^111^, this comes with the danger of blaming the victims of structural violence for its effects.

A final comment here relates to the quality of data regarding Covid-19 outcomes. Such was the speed at which the pandemic achieved great size, the data collection infrastructures were overwhelmed, and the quality of data describing its effects are not currently of a quality most would hope to have. This means that commentaries on effects on population health outcomes, especially those relying on quantitative data, should be treated with caution until better data is produced. This effect is most prominent in those commentaries undertaken early in the pandemic when the quality of data was at its worst. Furthermore, a number of those accounts authored in the early stages of the pandemic may have produced different conclusions if they were undertaken with a longer period of experience, evidence and context from which to draw.

## Discussion and Conclusions

Social factors have undoubtedly influenced the course and experiences of the Covid-19 pandemic. One important set of social factors is relationships – within and across societies. Social cohesion is a concept which might provide a useful tool in analysing and understanding the way relationships have operated and changed in the context of the pandemic. There is a body of literature which has made use of social cohesion and related concepts in this way. These use a range of constructions of social cohesion. This review categorised them into the following broad groups:

1. Those considering it a product of interpersonal relationships
2. Those claiming a reliance on sameness
3. Social cohesion as collective action and/or acting for the benefit of the collective
4. The accumulation of individual subjective perceptions or emotions relating to togetherness
5. The operation of structures of governance
6. Locally and/or culturally specific arrangements
7. Hybrid models

Some commentary centres on the effects of social cohesion on other objects or processes of interest and some on social cohesion itself during the pandemic. Of the former group, the following broad themes are present:

1. Cohesive groups or societies are generally said to see lower burdens of ill-health during the pandemic, depending on how social cohesion is constructed and the population under study.
2. Cohesive groups or societies are generally said to engage in better health-related practices in the context of Covid-19, though there are some tensions identified between the requirements of the larger collective and smaller groups.
3. Social cohesion is said to be a resource for resilience, emotional wellbeing and protection against clinically diagnosable mental states during the pandemic.
4. There are indications of an emergence of novel social groupings in relation to the demands of the pandemic and related policy and evidence of in-group / out-group dynamics.

Of the latter group, the following themes are present:

1. Changes to social cohesion claimed during the pandemic depend on the way it is constructed and the group(s) under investigation. Changes are distributed and experienced unequally.
2. There was a ‘rally round the flag’ moment early in the pandemic where many populations exhibited higher social cohesion by many different appraisals. This gave way to a return to type – or worse – toward the end of 2020.
3. Government policy prior to and during the pandemic has been of real importance to the operation of social cohesion ongoing.
4. There have been significant and fundamental changes to the practices around interpersonal relationships. These changes have not been distributed equally across populations.

Problems identified in the literature by and large reflect the wider and well-documented issues with the social cohesion concept and the diversity of forms it takes^33, 35, 106^. This makes comparison across the literature and the development of coherent thematic structures somewhat problematic. There is an over-reliance on long-established quantitative tools of measurement which are likely not entirely appropriate for such a new and complex situation. One of the major issues in this respect is the reduction of a complex and diverse arrangement of culturally-bound relationships to a limited number of closed-ended quantitative measures. There is a need for more detailed and in-depth theoretical and qualitative work and a focus on population groups of different sizes.

Much of what has been reported on here aligns with previous relevant scholarship on social cohesion. Interrelationships between social cohesion and health have been studied extensively, usually suggesting a positive association and a health promoting effect (e.g.^36, 112^). The importance of social cohesion in preparing for and responding to times of difficulty effectively has also been the topic of extensive scholarship, including violent conflict^113^, significant social change^114^, natural disasters^115, 116^ and pandemics^117, 118^. Moreover, the transmission of social cohesion to individual psychological and emotional resilience against distress is well studied (e.g.^119^) The situation-specific forms of bonding, collective action and support in response to moments of increased need (the “rally round the flag” effect^27^) has also been studied previously under the banner of social cohesion (e.g. ‘emergent social cohesion’^120^), such that scholars early in the Covid-19 pandemic engaged in hurried efforts to set out how this effect might be promoted and harnessed^121^.

However, some new knowledge has been produced by this literature. Obviously, some of this is in relation to Covid-19 itself which is, of course, itself new; but there has also been novel insight and development of existing ideas on how social cohesion may be conceptualised, its operation and important considerations therein. A good portion of the more notable insights are raised by qualitative work, which is well-represented in the body of work reviewed here and which is uncharacteristic of social cohesion scholarship in general. One of the more significant directions taken up by the literature dealing with social cohesion and the Covid-19 pandemic is the acknowledgement of a diversity of potentialities for social cohesion. These included the new arrangements of identities, group membership, relationships and manners of interaction demanded by the pandemic environment and policy responses^63, 64, 73, 102^. Positions maintaining that distinct locally and culturally bound arrangements of cohesion exist were also forwarded, taking up Green and Janmaat’s^110^ until now somewhat neglected invitation^76, 90, 99^. Commentaries also extended the understanding of the temporal nature of certain expressions of cohesion^27, 44, 60^.

Another area of interest where significant new ground was broken was the place of information and communication technology in the functioning of social relationships and therefore social cohesion, an area identified as neglected by Bayliss et al.^9^ in their 2019 review. Commentaries highlighted the importance of communications technology (and, of course, having possession of it and necessary skills to make use of it) for maintaining relationships over distance and therefore for the strengthening of cohesion^68, 83^; and thus by implication calling into question the usefulness of constructions of the concept which rely on face-to-face interaction. Also, importantly, scholarship drew attention to the technologically mediated processes of in/out group formation and political polarisation^54, 95, 98^.

This review has several limitations. Firstly, although as broad a net as possible was cast intentionally, the knowledge being produced is bounded by the limits of the search terms deployed. There are, for example, relevant literatures dealing with social capital (and not mentioning cohesion), communitas, ‘tight and loose’ cultures and more that are not captured by the singular focus on social cohesion.

Secondly, there are likely relevant works which were not held by any of the three libraries searched here and which have thus been omitted. Thirdly, as has been suggested in the findings section, during the period from which literature was obtained the situation and knowledge on it developed – and in many cases misunderstandings and errors were corrected – at great pace. This means that those published earlier in the pandemic did not have the benefit of the developments, context, corrections and hindsight that those published later did. This means that the knowledge being produced here is, at least in part, also subject to this issue. Fourthly, related to the third, data quality relating to Covid-19 (especially on morbidity and mortality and, again, especially from early in the pandemic) is still not of the standard one would wish to have to make robust claims. Finally, the great diversity of forms and manners in which social cohesion has been conceptualised, constructed and measured means that it is frequently difficult and problematic to make broad and sweeping statements on the body of literature. Indeed, as is common across social cohesion literatures, much of the scholarship uses similar nomenclature but is commenting upon and measuring a surprisingly diverse range of phenomena, experiences and subjectivities.

### Future Directions

Based on the above review and critique, there are a number of areas where future research may be focused to improve the body of knowledge. Firstly, there is a need for more engagement with theory within the social cohesion concept to sharpen it and make it more useful to understand the Covid-19 pandemic. There are some promising beginnings offered by those such as Schröder et al.^76^ which can be developed and put to use in developing the social cohesion concept itself, its components and ways of measurement that may aid a more complete understanding of the social world and its relationship with health. This will aid in ending an over-reliance on the more limited measures discussed. Following this, there is a need for more qualitative research in order to generate new knowledge on the way social cohesion may be understood and may operate in the context of Covid-19. Existing works such as that from Morgan et al.^83^ are an indication of the usefulness of detailed and in-depth qualitative work for understanding the unprecedented social conditions and relationships operating during the pandemic, and for the opportunity to develop the quantitative frameworks toward greater depth of understanding. Work in both of these arenas will assist both in improving the social cohesion concept and the social cohesion scholarship on the Covid-19 pandemic in relation to cultural specificity. The importance of this line of inquiry has been noted by Green and Janmaat^110^ for social cohesion generally and by Schröder et al.^76^ on the specific topic dealt with in this review, but work of this nature remains thin on the ground.

The literature in this body of work so far contains an over-representation of analyses conducted at the neighbourhood level. More work is needed to understand the operation of societies at the national level – the ongoing work conducted by Abrams and colleagues^59–62^ in the UK offer a good example of the value of such scholarship. There is also room for more analysis at the small-group level: only two works in the literature reviewed here attended to this^48, 65^. The operation of information and communication technology in respect of social cohesion also represents a promising avenue for future research offered by the work reviewed.

## Data Availability

Data from peer-reviewed journals

## Acknowledgements

Thanks are due to Drs. Peter Adams, Bruce Cohen and Peter Saxton for feedback, advice and editing of drafts

